# Predicting the future development of diabetic retinopathy using a deep learning algorithm for the analysis of non-invasive retinal imaging

**DOI:** 10.1101/2022.03.31.22272079

**Authors:** Yovel Rom, Rachelle Aviv, Sean Ianchulev, Zack Dvey-Aharon

## Abstract

**Aims:** Diabetic retinopathy (DR) is the most common cause of vision loss in the working age. This research aimed to develop a machine learning model which can predict the development of referrable DR from fundus imagery of otherwise healthy eyes.

**Methods:** Our researchers trained a machine learning algorithm on the EyePacs dataset, consisting of 156,363 fundus images. Referrable DR was defined as any level above mild on the International Clinical Diabetic Retinopathy scale.

**Results:** The algorithm achieved 0.81 Area Under Receiver Operating Curve (AUC) when averaging scores from multiple images on the task of predicting development of referrable DR, and 0.76 AUC when using a single image.

**Conclusion:** Our results suggest that risk of DR may be predicted from fundus photography alone. Prediction of personalized risk of DR may become key in treatment and contribute to patient compliance across the board, particularly when supported by further prospective research.

**Synopsis:** The deep learning algorithm our researchers developed was able to predict the development of referrable diabetic retinopathy in diabetic patients with otherwise healthy eyes with 0.81 AUC.

## Introduction

Diabetic retinopathy (DR) is a common retinal vascular complication of diabetes mellitus, which is characterized by retinal micro-aneurisms, hemorrhages, neovascularization, and edema in the retina.^1^ Diabetic retinopathy can advance to blindness, and is the leading cause of vision-loss at the working age. While over 80% of diabetics develop retinopathy of some degree after 20 years of the disease,^2^ more than 90% of the sight-threatening cases can be treated, if found early, in time to prevent loss of sight.^3^

Current public health guidelines for individuals with diabetes prescribe screening every 12-24 months for the presence of DR.^4,5^ Clinical studies have demonstrated that screening can lead to early detection and timely treatment which ultimately can prevent serious visual impairment and blindness.^6,7^ While retinal screening is essential for patients with diabetes, it requires a specialized eye exam which is often inaccessible for patients. A large percentages of individuals with diabetes forego screening retinal exams and present late in the course of the disease.^8–11^ Early intervention is the key to mitigation of DR risk factors and damage. As such, early detection is the most promising way of mitigating the damages of DR.

AI and machine learning have recently been successfully applied to the autonomous diagnosis of referrable (more than mild) DR. One FDA-approved AI system reported sensitivity of 87% and specificity of 90%.^12^ More recently, we reported results of a Pivotal FDA study with 93% sensitivity and 91% specificity for referrable DR on images obtained by a desktop device and 92% and 94% sensitivity and specificity, respectively, on images obtained by a portable camera.^13^ Additionally, we presented strong efficacy for DR detection using a portable camera on a separate dataset.^14^

Recent work has shown that otherwise “normal” fundus images can be informative and predictive when presented to a machine learning algorithm.^15–17^ AI algorithms can interpret subclinical information of the retinal anatomy and make predictions about diseases, even those unrelated to the eye - such as chronic kidney disease (CKD),^15^ diabetes,^16^ and cardiovascular risk factors.^17^ Furthermore, machine learning algorithms have been trained to predict gender information with high accuracy from mere fundus photography – something previously unattainable with the standard clinical exam.

While some work has been done on finding risk factors for DR, using patient data such as age, HbA1c levels, gender, duration of disease, and the like,^18,19^ clinicians are traditionally unable to predict the development of DR in patients. However, a previous article published findings of AUC 0.79 using a machine learning algorithm to predict DR development using fundus photography.^20^ These findings improved to 0.81 when combined with patient-specific information on risk factors. In this study we present a first-in-class machine learning algorithm which predicts the development of future DR from otherwise normal retinal anatomy. The current study improves on previous work by predicting the development of DR over a longer period of time, namely improving the prediction period from two to over three years, which may be clinically significant.

## Materials and Methods

### Dataset

We utilized a dataset compiled and provided by EyePACS (http://www.eyepacs.org), comprised of fundus retinal images and expert readings of said images. The data consisted of 156,363 images from 21,730 patients who visited the clinics at least twice between 2016 and 2021. 19.6% of visit pairs were ≤ 12 months apart, 55% were 12-24 months apart, 19.8% were 24-36 months apart, and 5.5% of visits were ≥ 36 months apart (Appendix A). Of the patients, 37% were male and 63% were female or other; mean age was 55 years old (Table 1). All images and data were de-identified according to the Health Insurance Portability and Accountability Act “Safe Harbor” before they were transferred to the researchers. Institutional Review Board exemption was obtained from the Sterling Independent Review Board.

**Table 1:** Key characteristics of the dataset.

|  | Training set | Validation set |
| --- | --- | --- |
| Number of patients | 19,531 | 2,133 |
| Number of images | 140,614 | 15,313 |
| Age: mean, years (s.d.) | 55.15 (10.68), n=19495 | 55.19 (10.38), n=2130 |
| Gender (% male) | 0.37, n=19102 | 0.37, n=2090 |
| HbA1c: mean, % (s.d.) | 7.98 (2.26), n=15672 | 7.99 (2.75), n=1703 |
| Disease duration: mean, years (s.d.) | 7.46 (6.44), n=18803 | 7.21 (6.20), n=2045 |
| Ethnicity | 62.1% Latin American, 11.4% ethnicity not specified, 9.1% African Descent, 7.4% Caucasian, 5.4% Asian, 2.8% Indian subcontinent origin, 1.1% Other, n=19076 | 61.4% Latin American, 10.7% ethnicity not specified, 9.7% African Descent, 7.8% Caucasian, 5.6% Asian, 3.1% Indian subcontinent origin, 1.1% Other, n=2086 |

The dataset contained up to 6 images per patient visit: one macula centered image, one disk centered image, and one centered image, per eye. Each eye was graded individually by an expert ophthalmologist for the presence and severity of diabetic retinopathy (DR). DR severity (none, mild, moderate, severe, or proliferative) was graded according to the International Clinical Diabetic Retinopathy scale.^21,22^

The image categorization in the current research was simplified to three severity categories by combining categories 3 to 5 into “more-than-mild DR”, as only these levels usually necessitate referral to an ophthalmologist and/or medical and surgical management.^23,24^

In order to prepare the dataset for model training and validation, each image was labeled by the maximal DR rating the patient was diagnosed with in a time period following the visit. Towards this purpose each patient visit was rated twice on the DR scale, once for each of the patient’s two eyes. Pairs were then created consisting of all possible pairings of each patient’s visits in a given time period. Values of the pairings were calculated by measuring the difference in DR ratings, and then taking the maximum value. Each time point (visit) was then assigned the highest value from the pairings in which that timepoint was the first, and each image was labeled by that value. Negative differences were disregarded, as the regression cause was unknown: true disease regression, clinical intervention, or misdiagnosis. Models were created for each of the chosen time periods.

For instance, a given patient has visited a clinic n times, once a year: v1, v2, …, vn. The cutoff for the given time period is set at two years, resulting in the following n-1 data points: v1 compared to v2 and v3 (taking the maximal difference), v2 compared to v3 and v4 (taking the maximal difference), etc. Further models using this patient’s data are also created, set at different time periods (three years, four years, and so on).

There were two reasons for choosing the maximal difference as the label. First, the main clinical value is in predicting whether a patient will develop DR, not in which eye it will be. Second, correlation found between the maximal right and left eye differences was relatively high (0.5), which indicates that difference between the eyes may well be incidental.

### Algorithm development

To evaluate the models’ performance, a random 10% of the patients were designated as the validation set and not used for the training of models. Of these, all images deemed fully gradable were used. Given that this same 10% were used as the validation set across tasks, this choice made the fair comparison of different models and timeframes easier.

In order to train the model, all the datapoints representing a progression were included, and a subset of the negative datapoints were included at a ratio of 2:1.

Models were trained on four different tasks:

- Progression amongst DR patients (mild to more-than-mild).
- Prediction of DR development (normal to any DR).
- Prediction of clinically significant DR (from non-referrable to referrable DR).
- General progression (progress): any change for the worse in the DR condition.

The hyper-parameters for the model training were chosen beforehand, and not changed, to prevent over-fitting.

### Risk factor predictive value

For 80% of the patients in the dataset, HbA1c level was recorded by the clinic, and disease duration was recorded for 98% of patients. HbA1c level and disease duration were treated as risk level scores. AUC was then calculated in order to rate predictive value for each of the scores, which were then compared to the predictive value of the model on each task (table 3).

## Results

### Transitional Retinopathy Results

The calculated baseline transition odds between different DR levels are displayed in Table 2 (for a more detailed table see appendix B). This is observational data, as regression- and progression-related factors are unknown; regression may have been caused by clinical intervention, and progression may be understated due to patients who experienced vision loss and therefore did not return for subsequent visits.

**Table 2:**
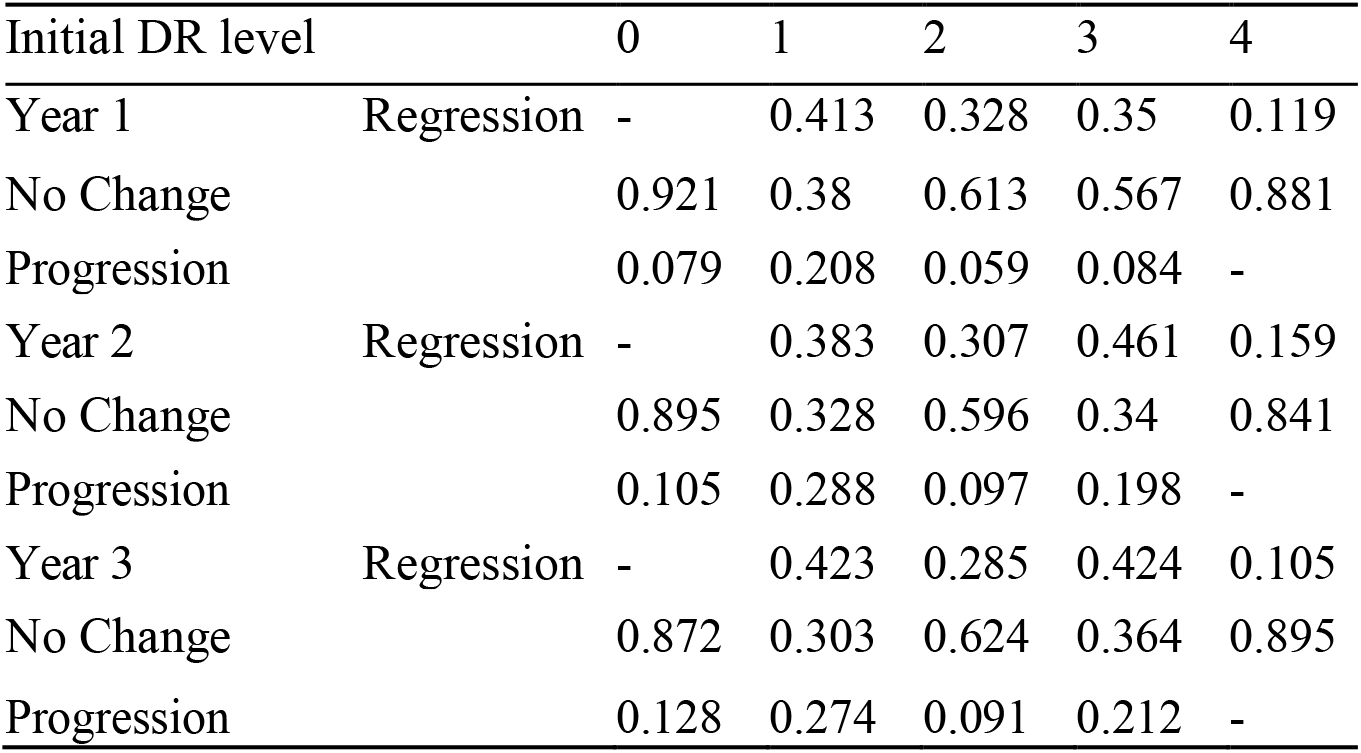
Disease progression was calculated between any given visit and the visit immediately following. “Regression” is defined as the patient’s recorded DR level being lower on the second visit, “No change” is defined as recorded DR levels being the same between visits, and “Progression” is defined as DR levels being higher.

### Prediction Results

The model’s performance in determining the risk of mild DR becoming more-than-mild DR is comparable to risk factor-based prediction (area under the receiver operating curve (AUC) 0.65 vs. 0.66, respectively). For the other tasks more images were available, (appendix J) and performance improved significantly. The results improved still by using multiple images per patient and averaging the resulting score. The model scored best on the task of prediction of clinically significant DR, with the aggregated score resulting in AUC 0.81 (CI 95% 0.77-0.84) (Table 3. Additional timeframes and ROC curves available in appendices G and H). As the HbA1c levels and disease duration was not available for all patients, model scores were also calculated for the subsets of patients who had those scores, resulting in effectively the same scores (appendix M). The Pearson correlation between our model’s score and HbA1c levels was 0.12 (CI 95% 0.06-0.21). Correlation with disease duration was 0.21 (CI 95% 0.22-0.30).

**Table 3:** The models’ score, in AUC, in predicting DR within two years. In the parentheses 95% CI. See supplemental for additional timeframes.

|  | Image Prediction (by algorithm) | Patient Prediction (by algorithm) | HbA1c-based prediction | Disease Duration-based prediction |
| --- | --- | --- | --- | --- |
| Mild DR to mtmDR | 0.63 (0.56, 0.70) | **0.65 (0.57, 0.73)** | **0.66 (0.62, 0.71)** | 0.50 (0.46, 0.54) |
| No DR to mtmDR | 0.67 (0.64, 0.70) | **0.71 (0.68, 0.74)** | 0.65 (0.63, 0.67) | 0.60 (0.59, 0.62) |
| mtmDR- to mtmDR+ | 0.75 (0.72, 0.78) | **0.81 (0.77, 0.84)** | 0.70 (0.68, 0.72) | 0.63 (0.62, 0.65) |
| Any DR Progression | 0.71 (0.68, 0.73) | **0.75 (0.72, 0.78)** | 0.67 (0.66, 0.69) | 0.61 (0.60, 0.62) |

To further analyze the model’s prediction value in that task, the empirical risk as a function of the model’s score was investigated (Figure 1). When the model was trained to predict the transition to more-than-mild DR, the top 5% patients highest scored by the model were at 54% risk of getting DR, while the baseline odds in the validation set were 10% - almost a 5-fold increase.

**Figure 1:**
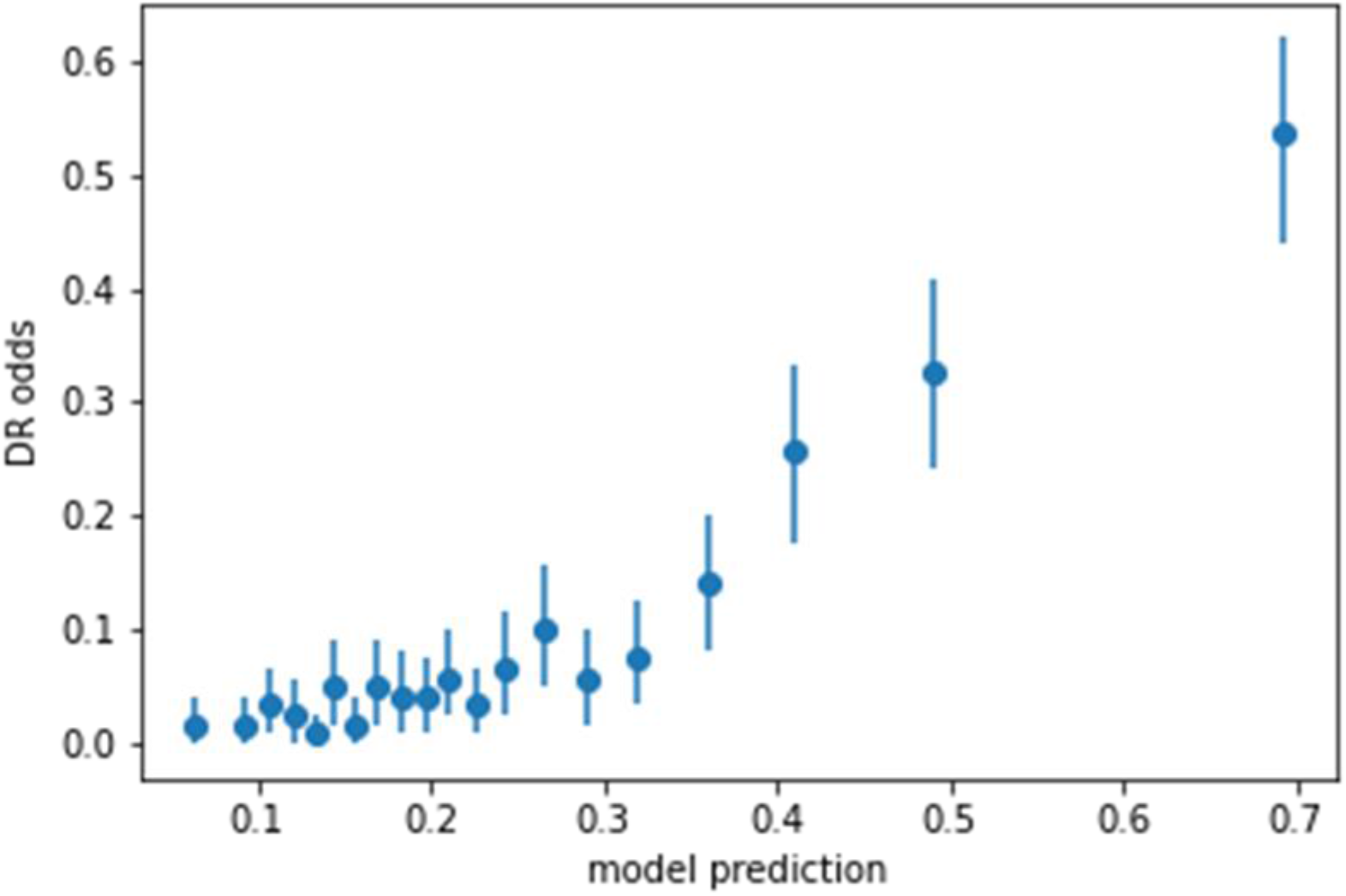
the odds of the patient from a representative sample being diagnosed with clinically significant DR in two years, as a function of the model’s score. Each dot represents 5% of the patients. Additional figures in appendix K

**Figure 2:**
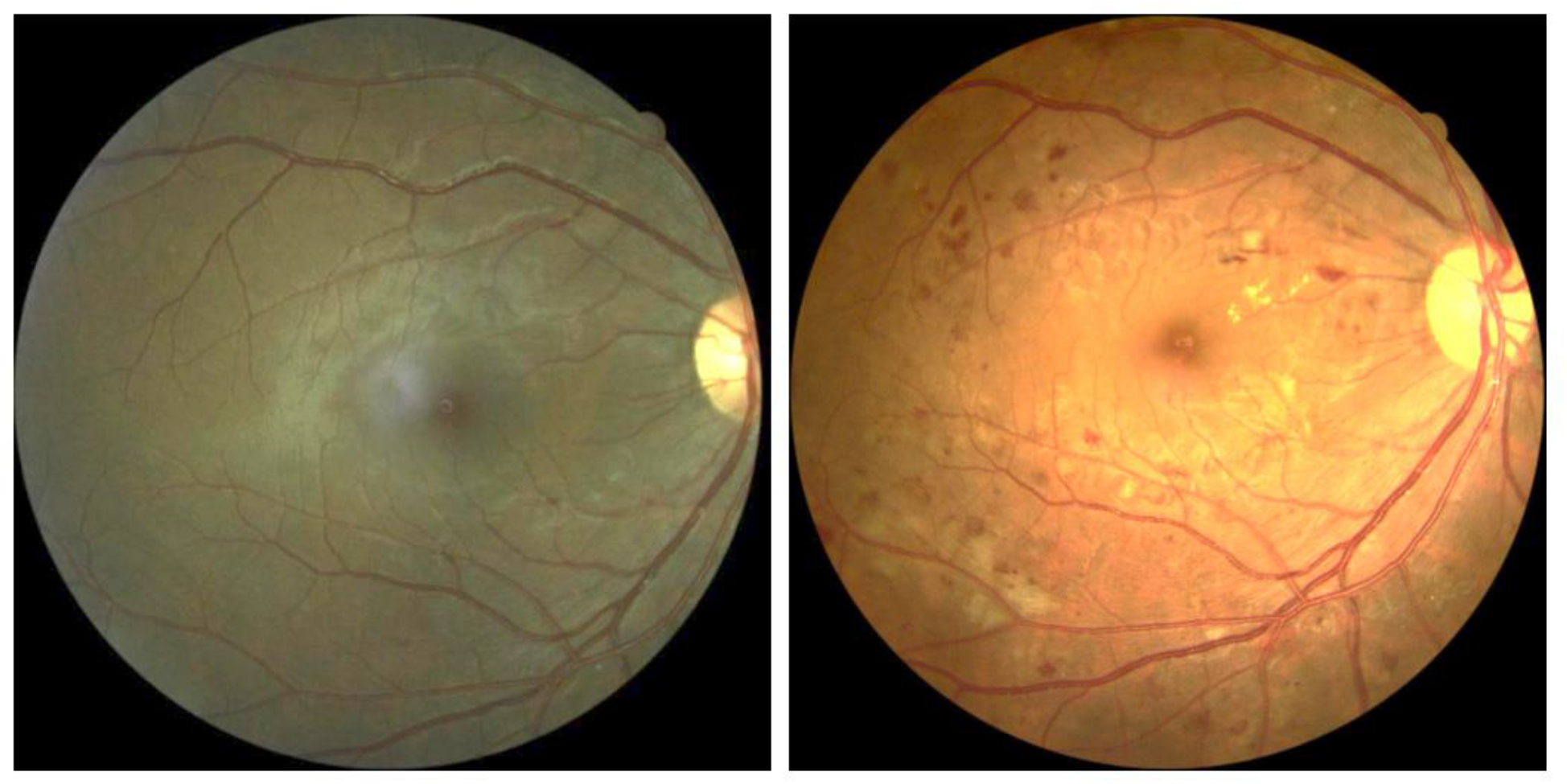
Left: the right eye at the first timepoint, rated healthy by a human expert. Right: the same eye one year later, with severe DR. The model score for both healthy eyes was in the top 2% of severity, implying more than fivefold the baseline risk of developing more than mild DR in the following two years. One year later, the patient was diagnosed with severe and proliferative DR in the right and left eyes respectively.

In order to further analyze the model’s performance as a function of time, sets of images were split into four groups based on time elapsed between first and second visits and imaging (Table 4). In order to analyze the relation between the model’s assigned scores and the severity of developed DR, the scores assigned by the model were averaged and compared across different subgroups; normal patients who were diagnosed with DR up to two years after initial images were taken were sorted into subgroups based on DR severity. The groups of mild, moderate, and severe DR were score averaged at 0.36, 0.43, and 0.46 respectively (Figure 4).

**Table 4:** Comparison of model performance as a function of time. In the parentheses 95% CI.

| Prediction time (months) | Image Prediction (by algorithm) | Patient Prediction (by algorithm) |
| --- | --- | --- |
| Up to 15 | 0.73 (0.68, 0.78) | 0.79 (0.74, 0.84) |
| 16-24 | 0.76 (0.71, 0.80) | 0.80 (0.76, 0.84) |
| 25-36 | 0.69 (0.63, 0.74) | 0.75 (0.68, 0.80) |
| 36+ | 0.82 (0.73, 0.90) | 0.88 (0.67, 0.96) |

Additionally, the model’s predictive effectiveness regarding the more rapid development of DR was analyzed. Assigned scores were averaged and compared across subgroups organized by time elapsed between initial imaging and diagnosis: one, two, three, and four or more years. It was hypothesized that the healthier the eye appeared the more time would elapse between initial imaging and diagnosis. As expected, the model’s score declined in congruence with time passed before diagnosis (see Figure 3).

When compared with results from Bora et al., our model shows an improvement in performance from 0.79 AUC to 0.82,^20^ as well as the ability to predict the development of DR across a longer timeframe, up to five years. When additional relevant patient metadata was utilized, the former article improved results to 0.81 AUC. Given the lack of metadata in the dataset utilized for this research, results for this model may improve accordingly if validated on other datasets.

**Figure 3:**
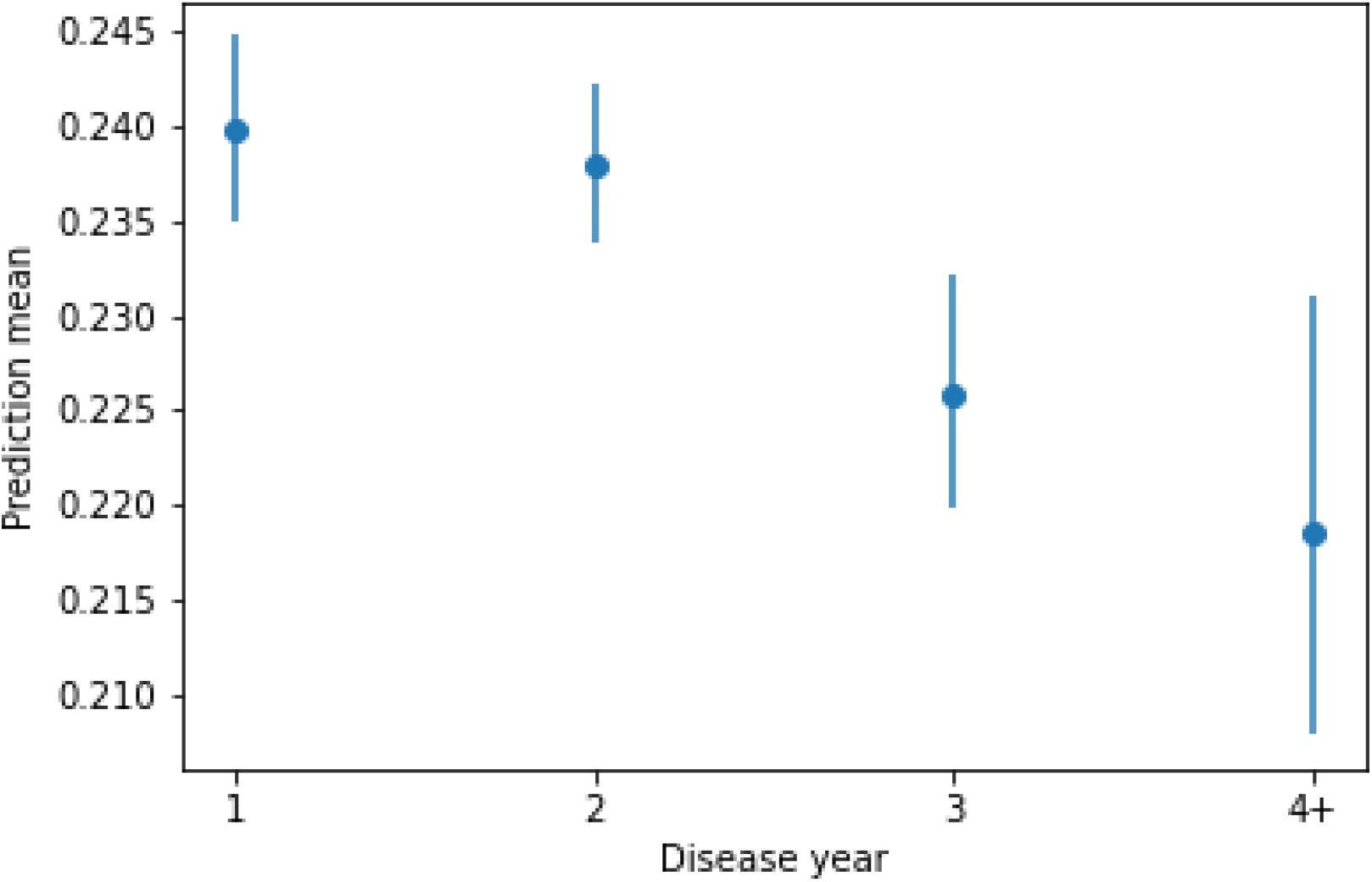
The model’s mean score as a function of how many years after the visit DR was diagnosed. As expected, model average score decreases in accordance with time elapsed between first visit and diagnosis

**Figure 4:**
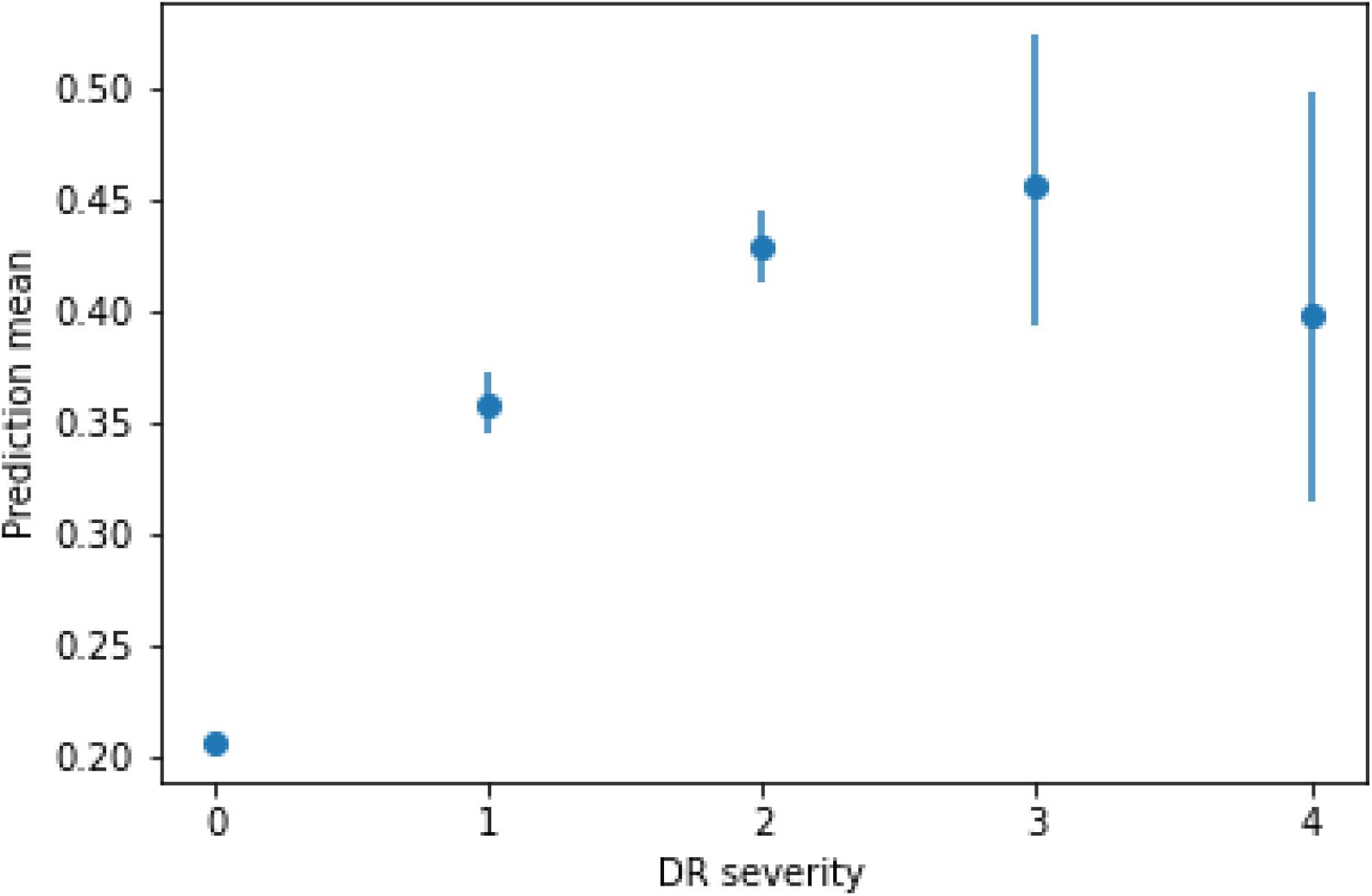
The model’s score as a function of the final diagnosed severity of the DR. The model is more confident of the future occurrence of DR if it’s more severe.

## Discussion

The aim of this research was to develop a method of predicting the chances of future development of DR, before detection methods can even be applied. Given the relatively high prevalence of DR,^2^ the difficulty in permanently reversing retinal damage,^25^ and current patient non-compliance,^8–11^ prevention and not only treatment of DR is crucial to health outcomes. As such, prediction of individual DR risk may become a key element. Currently, to the best of our knowledge, there are two methods of doing this: risk factor based, which is of limited predictive clinical utility,^18,19^ and that developed by Bora et al.^20^ which utilizes both deep learning and risk factors for optimal results.

The current research was conducted using convolutional neural networks, a standard state of the art computer vision algorithm. Such algorithms have been reliably incorporated in multiple medical fields, such as ophthalmology,^26^ radiology,^27^ endocrinology,^28^ and others.^29^ The algorithms presented in this work, which are easily implemented and display promising performance, may carry widespread implications related to better DR prediction. For instance, given the knowledge that some patients are at very low risk of developing DR, screening may be able to be feasibly reduced according to individual risk levels, reducing strain placed on both patients and medical staff. Furthermore, in high-risk cases which have not yet manifested, patients may be forewarned of impending risk, increasing chances of mitigation and prevention through diabetes management. Bora et al. previously demonstrated the ability to predict the development of DR within two years.^20^ The current research is able to predict development within over three, which may have practical and clinical implications. Due to clinical guidelines recommending routine patient checkups every one to two years, the increase in predictive time may decrease the number of screenings required for patients, affording longer periods of time between necessary checkups.

A lack of patient education regarding the risks of diabetes, and DR specifically, has been cited as a contributing factor in patient non-compliance.^8^ Furthermore, patients may not attend screenings due to belief that they do not require retinal examinations or treatment as their vision is too good, or their diabetes is too mild to be relevant. The ability to concretely discuss personal risk levels with the patient may do much to mitigate these beliefs, contributing to higher compliance. Improved patient compliance in terms of DR may also improve compliance in terms of general diabetes management, bettering patient outcomes across the board.

One limitation of the current research is that the transition odds are observatory, rather than experimental. As such, there is the possibility of an under-statement of risk, given that blinded people likely did not continue to return for checkups. Odds of regression may similarly be over-stated, as regression factors are unknown and regression may have been caused by surgical or medical intervention.

Recommendations for future research include studies on how to incorporate the model into usual diabetes standards of care, in order to better mitigate and prevent DR. As the model’s score was not strongly correlated to risk factor score, there may be added value in including metadata on levels of previously recognized risk factors among patients in order to improve predictive value, as demonstrated in previous studies.^20^ Additionally, there is great value in examining whether use of this algorithm does, in fact, improve patient compliance. This model may also contribute to future research of DR risk factors and prevention, as at-risk patients with previously unknown risk factors may become recognizable, allowing for a more holistic understanding of contributing influences, both biological and behavioral.

## Supporting information

article supplements

## Data Availability

All data produced in the present study is proprietary. Anonymized patient IDs are available upon request to the authors.

## References

1 Cheung N, Mitchell P, Wong TY. Diabetic retinopathy. Lancet 2010;376: 124–36.

2 Kertes PJ, Johnson TM. Evidence-based eye care. Lippincott Williams & Wilkins, 2007.

3 Tapp RJ, Shaw JE, Harper CA, et al. The prevalence of and factors associated with diabetic retinopathy in the Australian population. Diabetes care 2003;26: 1731–7.

4 American Diabetes Association Professional Practice Committee, Draznin B, Aroda VR, et al. Retinopathy, Neuropathy, and Foot Care: Standards of Medical Care in Diabetes-2022. Diabetes Care 2022;45: S185–94.

5 World Health Organization. Regional Office for Europe. Diabetic retinopathy screening: a short guide: increase effectiveness, maximize benefits and minimize harm. World Health Organization. Regional Office for Europe, 2020 https://apps.who.int/iris/handle/10665/336660 (accessed Feb 27, 2022).

6 Rohan TE, Frost CD, Wald NJ. Prevention of blindness by screening for diabetic retinopathy: a quantitative assessment. BMJ 1989;299: 1198–201.

7 Stefánsson E, Bek T, Porta M, Larsen N, Kristinsson JK, Agardh E. Screening and prevention of diabetic blindness. Acta Ophthalmol Scand 2000;78: 374–85.

8 Lewis K. Improving patient compliance with diabetic retinopathy screening and treatment. Community Eye Health 2015;28: 68–9.

9 National Institute for Health and Care Excellence (NICE). Type 2 diabetes in adults: management. NICE https://www.nice.org.uk/guidance/ng28 (accessed Feb 27, 2022).

10 Paz SH, Varma R, Klein R, Wu J, Azen SP, Los Angeles Latino Eye Study Group. Noncompliance with vision care guidelines in Latinos with type 2 diabetes mellitus: the Los Angeles Latino Eye Study. Ophthalmology 2006;113: 1372–7.

11 Saadine JB, Fong DS, Yao J. Factors associated with follow-up eye examinations among persons with diabetes. Retina 2008;28: 195–200.

12 Abràmoff MD, Lavin PT, Birch M, Shah N, Folk JC. Pivotal trial of an autonomous AI-based diagnostic system for detection of diabetic retinopathy in primary care offices. npj Digital Med 2018;1: 1–8.

13 AEYE. AEYE Health Reports Pivotal Clinical Trial Results of its AI Algorithm for the Autonomous Screening and Detection of More-Than-Mild Diabetic Retinopathy. https://www.prnewswire.com/news-releases/aeye-health-reports-pivotal-clinical-trial-results-of-its-ai-algorithm-for-the-autonomous-screening-and-detection-of-more-than-mild-diabetic-retinopathy-301476299.html (accessed Feb 28, 2022).

14 Dvey-Aharon Z, Huhtinen P. Screening for Diabetic Retinopathy in Endocrinology Clinics by Using Handheld Cameras and Applying Artificial Intelligence Algorithms. J Endocr Soc 2021;5: A419–20.

15 Sabanayagam C, Xu D, Ting DSW, et al. A deep learning algorithm to detect chronic kidney disease from retinal photographs in community-based populations. The Lancet Digital Health 2020;2: e295–302.

16 Mitani A, Hammel N, Liu Y. Retinal detection of kidney disease and diabetes. Nat Biomed Eng 2021;5: 487–9.

17 Poplin R, Varadarajan AV, Blumer K, et al. Prediction of cardiovascular risk factors from retinal fundus photographs via deep learning. Nat Biomed Eng 2018;2: 158–64.

18 Magliah SF, Bardisi W, Al Attah M, Khorsheed MM. The prevalence and risk factors of diabetic retinopathy in selected primary care centers during the 3-year screening intervals. J Family Med Prim Care 2018;7: 975–81.

19 Sheu S-J, Liu N-C, Ger L-P, et al. High HbA1c level was the most important factor associated with prevalence of diabetic retinopathy in Taiwanese type II diabetic patients with a fixed duration. Graefes Arch Clin Exp Ophthalmol 2013;251: 2087–92.

20 Bora A, Balasubramanian S, Babenko B, et al. Predicting the risk of developing diabetic retinopathy using deep learning. The Lancet Digital Health 2021;3: e10–9.

21 Haneda S, Yamashita H. International clinical diabetic retinopathy disease severity scale. Nihon rinsho Japanese journal of clinical medicine 2010;68: 228–35.

22 Gulshan V, Peng L, Coram M, et al. Development and Validation of a Deep Learning Algorithm for Detection of Diabetic Retinopathy in Retinal Fundus Photographs. JAMA 2016;316: 2402–10.

23 Flaxel CJ, Adelman RA, Bailey ST, et al. Diabetic Retinopathy Preferred Practice Pattern®. Ophthalmology 2020;127: P66–145.

24 American Optometric Association. Eye Care of the Patient with Diabetes Mellitus. US, 2019 https://www.aoa.org/AOA/Documents/Practice%20Management/Clinical%20Guidelines/EBO%20Guidelines/Eye%20Care%20of%20the%20Patient%20with%20Diabetes%20Mellitus%2C%20Second%20Edition.pdf.

25 Wykoff CC, Eichenbaum DA, Roth DB, Hill L, Fung AE, Haskova Z. Ranibizumab Induces Regression of Diabetic Retinopathy in Most Patients at High Risk of Progression to Proliferative Diabetic Retinopathy. Ophthalmology Retina 2018;2: 997–1009.

26 Food and Drug Administration (FDA). DE NOVO CLASSIFICATION REQUEST FOR IDX-DR. Coralville, IA, 2018 https://www.accessdata.fda.gov/cdrh_docs/reviews/DEN180001.pdf (accessed Feb 28, 2022).

27 Food and Drug Administration (FDA). EVALUATION OF AUTOMATIC CLASS III DESIGNATION FOR OSTEODETECT. Coralville, IA, 2018 https://www.accessdata.fda.gov/cdrh_docs/reviews/DEN180005.pdf (accessed Feb 28, 2022).

28 Food and Drug Administration (FDA). P160007 Approval Order. Coralville, IA, 2018 https://www.accessdata.fda.gov/cdrh_docs/pdf16/P160007A.pdf (accessed Feb 28, 2022).

29 Benjamens S, Dhunnoo P, Meskó B. The state of artificial intelligence-based FDA-approved medical devices and algorithms: an online database. npj Digit Med 2020;3: 1–8.

