## Supplementary material for "Predicting the future development of diabetic retinopathy using a deep learning algorithm for the analysis of non-invasive retinal imaging": article supplements

### 5. Appendices

## 6.1 A

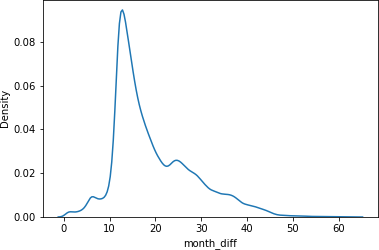

## 6.2 B

DR progression, by initial stage. Regression includes recovery by surgery.

| One year: |  |  |  |  |  |  |
| --- | --- | --- | --- | --- | --- | --- |
|  |  | 0 | 1 | 2 | 3 | 4 |
|  | -4 | - | - | - | - | 0.0042 |
|  | -3 | - | - | - | 0.0083 | 0.0084 |
|  | -2 | - | - | 0.199 | 0 | 0.0816 |
|  | -1 | - | 0.4127 | 0.129 | 0.3412 | 0.0249 |
|  | 0 | 0.9209 | 0.3798 | 0.6129 | 0.5667 | 0.8809 |
|  | 1 | 0.044 | 0.2045 | 0.0462 | 0.0838 | - |
|  | 2 | 0.0343 | 0.0026 | 0.0129 | - | - |
|  | 3 | 0.0004 | 0.0004 | - | - | - |
|  | 4 | 0.0004 | - | - | - | - |
| Two years: |  |  |  |  |  |  |
|  |  | 0 | 1 | 2 | 3 | 4 |
|  | -4 | - | - | - | - | 0.0111 |
|  | -3 | - | - | - | 0.0175 | 0.0148 |
|  | -2 | - | - | 0.1683 | 0.0175 | 0.0888 |
|  | -1 | - | 0.3834 | 0.1391 | 0.4261 | 0.0444 |
|  | 0 | 0.8951 | 0.3284 | 0.596 | 0.3405 | 0.8409 |
|  | 1 | 0.0576 | 0.2784 | 0.0667 | 0.1984 | - |
|  | 2 | 0.0461 | 0.0058 | 0.03 | - | - |
|  | 3 | 0.0008 | 0.0039 | - | - | - |
|  | 4 | 0.0003 | - | - | - | - |
| Three years: |  |  |  |  |  |  |
|  |  | 0 | 1 | 2 | 3 | 4 |
|  | -4 | - | - | - | - | 0 |
|  | -3 | - | - | - | 0.0909 | 0 |
|  | -2 | - | - | 0.1725 | 0.0909 | 0.1053 |
|  | -1 | - | 0.4229 | 0.1128 | 0.2424 | 0 |
|  | 0 | 0.8719 | 0.303 | 0.6241 | 0.3636 | 0.8947 |
|  | 1 | 0.0746 | 0.2622 | 0.042 | 0.2121 | - |
|  | 2 | 0.0504 | 0.0119 | 0.0485 | - | - |
|  | 3 | 0.0023 | 0 | - | - | - |
|  | 4 | 0.0009 | - | - | - | - |

## 6.3 C

The images in the original dataset were taken using several cameras. The exact division is shown in the following table:

|  | Camera Name | Number of Images |
| --- | --- | --- |
|  | Cannon CR2 | 263,815 |
|  | Centervue DRS | 145,257 |
|  | Crystalvue | 8,092 |
|  | Topcon NW400 | 367,854 |

| **6.4 D** |
| --- |
| HbA1c distribution: |

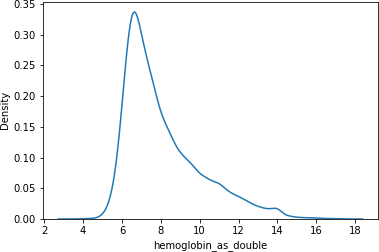

There were 20 patients with HbA1C values ranging from 56.2 to 1132 who were excluded from this figure

## 6.5 E

DR odds as a function of hemoglobin, first year:

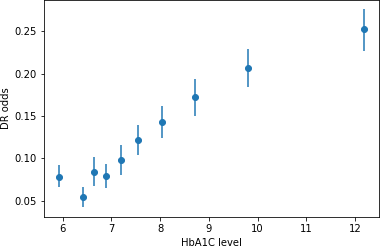

DR odds as a function of HbA1c, second year:

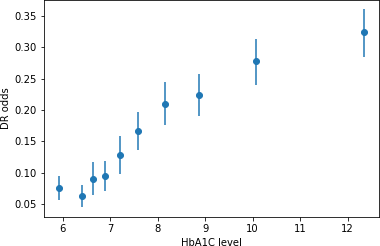

DR odds as a function of HbA1c, third year:

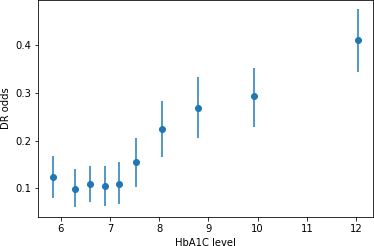

|  | **6.6 F**  Disease progression was calculated between any given visit and the visit immediately following. “Regression” is defined as the patient’s recorded DR level being lower on the second visit, “No change” is defined as recorded DR levels being the same between visits, and “Progression” is defined as DR levels being higher.  First year: | | | | |
| --- | --- | --- | --- | --- | --- |
|  |  | |  | specific image | specific patient |
|  |  | | mild_to_severe | 0.576 (0.515, 0.634) | 0.618 (0.495, 0.731) |
|  |  | | healthy_to_severe | 0.641 (0.621, 0.661) | 0.677 (0.626, 0.722) |
|  |  | | to_severe | 0.734 (0.710, 0.756) | 0.789 (0.738, 0.831) |
|  |  | | progress | 0.670 (0.652, 0.688) | 0.711 (0.666, 0.750) |
|  | Second year: | |  |  |  |
|  |  | |  | specific image | specific patient |
|  |  | | mild_to_severe | 0.630 (0.590, 0.670) | 0.653 (0.572, 0.728) |
|  |  | | healthy_to_severe | 0.667 (0.652, 0.681) | 0.708 (0.677, 0.738) |
|  |  | | to_severe | 0.746 (0.729, 0.761) | 0.807 (0.773, 0.837) |
|  | | | progress | 0.703 (0.690, 0.716) | 0.750 (0.722, 0.777) |
|  | | Three years: |  |  |  |
|  | |  |  | specific image | specific patient |
|  | |  | mild_to_severe | 0.611 (0.572, 0.648) | 0.635 (0.559, 0.705) |
|  | |  | healthy_to_severe | 0.667 (0.654, 0.680) | 0.712 (0.683, 0.739) |
|  | |  | to_severe | 0.725 (0.711, 0.740) | 0.779 (0.749, 0.807) |
|  | |  | progress | 0.710 (0.699, 0.721) | 0.753 (0.727, 0.776) |

|  | All images: |  |  |  |
| --- | --- | --- | --- | --- |
|  |  |  | specific image | specific patient |
|  |  | mild_to_severe | 0.603 (0.566, 0.641) | 0.629 (0.554, 0.697) |
|  |  | healthy_to_severe | 0.669 (0.656, 0.682) | 0.711 (0.683, 0.737) |
|  |  | to_severe | 0.729 (0.714, 0.743) | 0.785 (0.754, 0.811) |
|  |  | progress | 0.705 (0.694, 0.716) | 0.754 (0.729, 0.777) |

## 6.7 G

ROC curves of prediction by image.

Mild to more than mild:

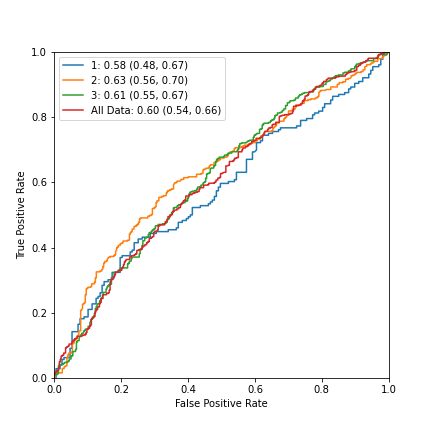

Normal to more than mild:

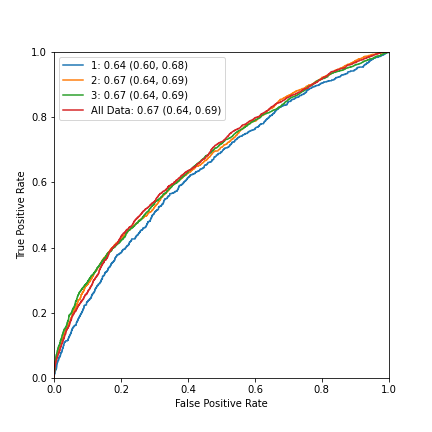

Non referrable to referrable:

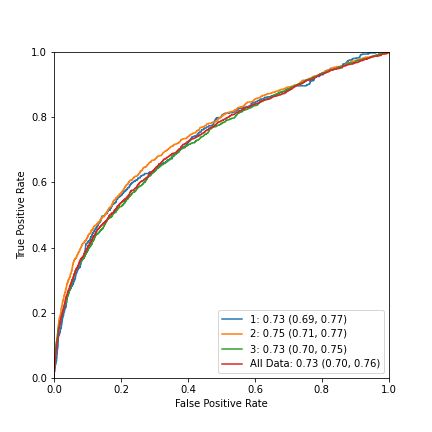

Any progression:

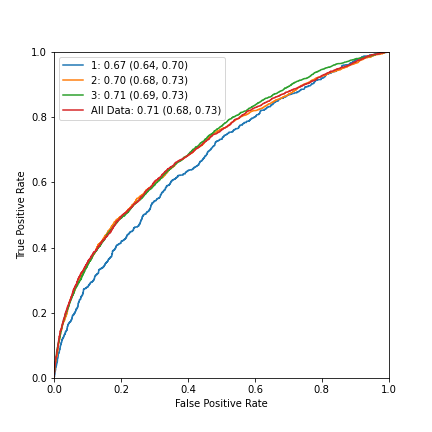

## 6.8 H

ROC curves of prediction by patient.

Mild to more than mild:

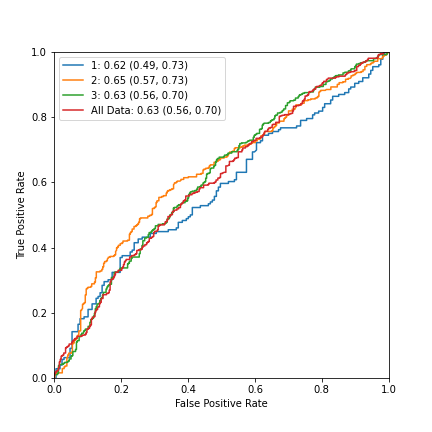

Normal to more than mild:

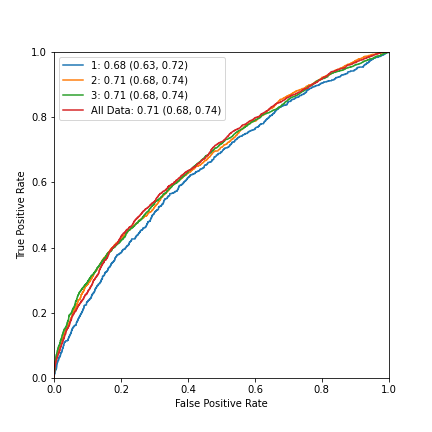

Non referrable to referrable:

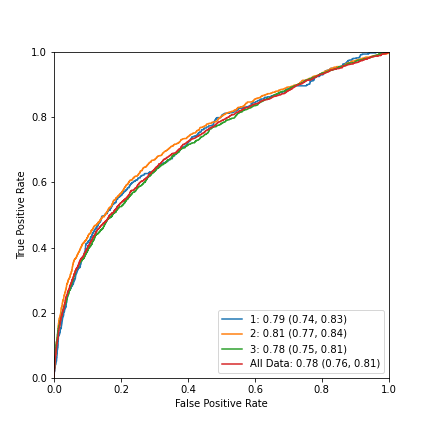

Any progression:

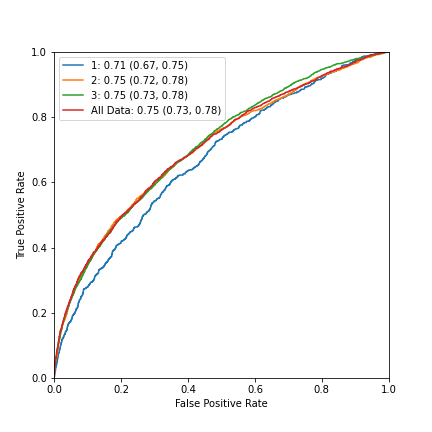

## 6.9 I

Demographic characteristics of datasets with different timeframes:

First year:

|  | **Training set** | **Validation set** |
| --- | --- | --- |
| **Number of patients** | 8,947 | 968 |
| **Number of images** | 59,405 | 6,548 |
| **Age: mean, years (s.d.)** | 55.66 (10.52), n=8928 | 55.88 (10.17), n=966 |
| **Gender (% male)** | 0.37, n=8753 | 0.38, n=952 |
| **HbA1c: mean, % (s.d.)** | 7.87 (1.87), n=7159 | 7.93 (2.78), n=766 |
| **Disease duration: mean, years (s.d.)** | 7.62 (6.45), n=8640 | 7.35 (6.05), n=922 |
| **Ethnicity** | 62.6% Latin American, 11.6% ethnicity not specified, 8.7% African Descent, 6.4% Caucasian, 5.8% Asian, 3.3% Indian subcontinent origin, n=8740 | 62.8% Latin American, 11.1% ethnicity not specified, 8.8% African Descent, 5.9% Caucasian, 5.5% Asian, 3.9% Indian subcontinent origin, 1.4% Other, n=947 |

Second year:

|  | **Training set** | **Validation set** |
| --- | --- | --- |
| **Number of patients** | 17,161 | 1,896 |
| **Number of images** | 124,053 | 13,716 |
| **Age: mean, years (s.d.)** | 55.26 (10.65), n=17126 | 55.52 (10.41), n=1892 |
| **Gender (% male)** | 0.37, n=16783 | 0.36, n=1856 |
| **HbA1c: mean, % (s.d.)** | 7.96 (2.16), n=13845 | 7.95 (2.42), n=1515 |
| **Disease duration: mean, years (s.d.)** | 7.47 (6.43), n=16514 | 7.29 (6.27), n=1814 |
| **Ethnicity** | 62.8% Latin American, 11.4% ethnicity not specified, 8.7% African Descent, 7.2% Caucasian, 5.5% Asian, 2.7% Indian subcontinent origin, 1.1% Other, n=16753 | 61.5% Latin American, 10.8% ethnicity not specified, 9.1% African Descent, 7.8% Caucasian, 5.9% Asian, 3.2% Indian subcontinent origin, 1.2% Other, n=1856 |

Third year:

|  | **Training set** | **Validation set** |
| --- | --- | --- |
| **Number of patients** | 19,172 | 2,152 |
| **Number of images** | 138,333 | 15,447 |
| **Age: mean, years (s.d.)** | 55.19 (10.65), n=19136 | 55.35 (10.41), n=2148 |
| **Gender (% male)** | 0.37, n=18748 | 0.37, n=2109 |
| **HbA1c: mean, % (s.d.)** | 7.97 (2.26), n=15375 | 7.99 (2.76), n=1716 |
| **Disease duration: mean, years (s.d.)** | 7.46 (6.45), n=18448 | 7.20 (6.21), n=2061 |
| **Ethnicity** | 62.2% Latin American, 11.4% ethnicity not specified, 9.0% African Descent, 7.4% Caucasian, 5.5% Asian, 2.8% Indian subcontinent origin, 1.1% Other, n=18719 | 61.2% Latin American, 10.8% ethnicity not specified, 9.7% African Descent, 7.8% Caucasian, 5.8% Asian, 3.1% Indian subcontinent origin, 1.1% Other, n=2100 |

All data:

|  | **Training set** | **Validation set** |
| --- | --- | --- |
| **Number of patients** | 19,531 | 2,199 |
| **Number of images** | 140,614 | 15,749 |
| **Age: mean, years (s.d.)** | 55.15 (10.68), n=19495 | 55.34 (10.40), n=2195 |
| **Gender (% male)** | 0.37, n=19102 | 0.37, n=2155 |
| **HbA1c: mean, % (s.d.)** | 7.98 (2.26), n=15672 | 7.99 (2.75), n=1756 |
| **Disease duration: mean, years (s.d.)** | 7.46 (6.44), n=18803 | 7.21 (6.20), n=2108 |
| **Ethnicity** | 62.1% Latin American, 11.4% ethnicity not specified, 9.1% African Descent, 7.4% Caucasian, 5.4% Asian, 2.8% Indian subcontinent origin, 1.1% Other, n=19076 | 61.4% Latin American, 10.7% ethnicity not specified, 9.7% African Descent, 7.8% Caucasian, 5.6% Asian, 3.1% Indian subcontinent origin, 1.1% Other, n=2147 |

## 6.10 J

Table describing the sizes of datasets. First number is number of images in the development set, second is validation set.

|  | Mild DR to mtmDR | No DR to mtmDR | mtmDR- to mtmDR+ | Any DR Progression |
| --- | --- | --- | --- | --- |
| **1** | 3415, 343 | 55990, 6205 | 59405, 6548 | 59405, 6548 |
| **2** | 7011, 745 | 117042, 12971 | 124053, 13716 | 124053, 13716 |
| **3** | 8121, 853 | 130212, 14594 | 138333, 15447 | 138333, 15447 |
| **4** | 8351, 885 | 132263, 14864 | 140614, 15749 | 140614, 15749 |

## 6.11 K

The empirical risk as a function of the model’s score was investigated. The following four plots show the empirical risk as a function of model score per image for different tasks. The four following plots show the same per patient, by averaging the scores of all the images taken during one visit.

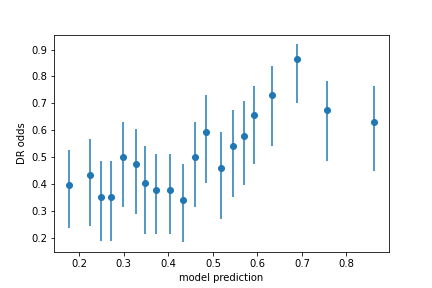

*Figure 5: Empiric risk of mild DR to mtmDR as a function of model score per image*

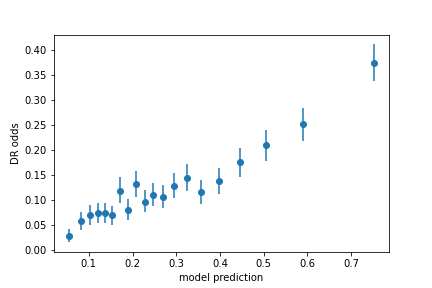

*Figure 6: Empiric risk of no DR to mtmDR as a function of model score per image*

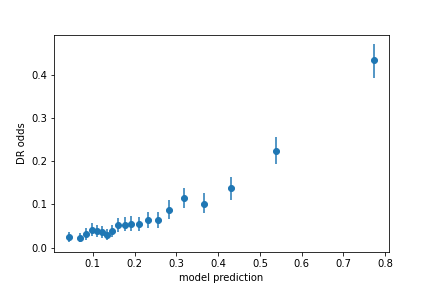

*Figure 7: Empiric risk of mtmDR- to mtmDR+ as a function of model score per image*

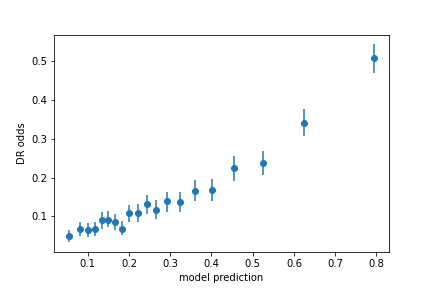

*Figure 8: Empiric risk of any DR progression as a function of model score per image*

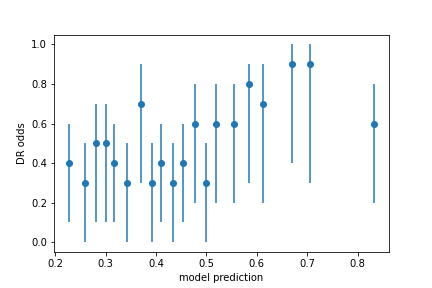

*Figure 9: Empiric risk of mild DR to mtmDR as a function of model score per patient*

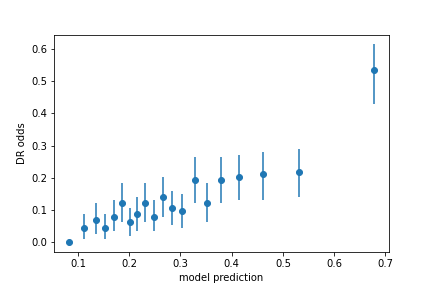

*Figure 10: Empiric risk of no DR to mtmDR as a function of model score per patient*

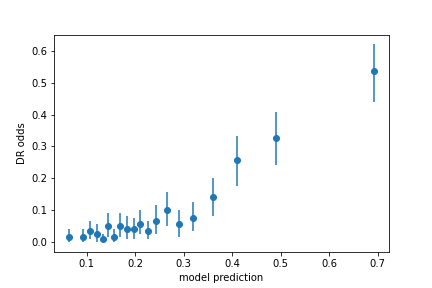

*Figure 11: Empiric risk of mtmDR- to mtmDR+ as a function of model score per patient*

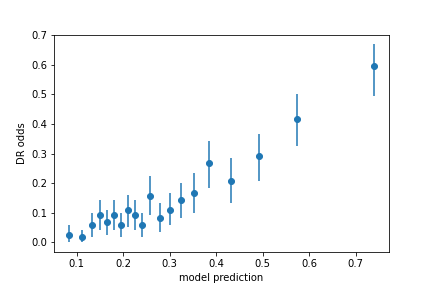

*Figure 12: Empiric risk of any DR progression as a function of model score per patient*

## 6.12 L

A figure describing our dataset creation process.

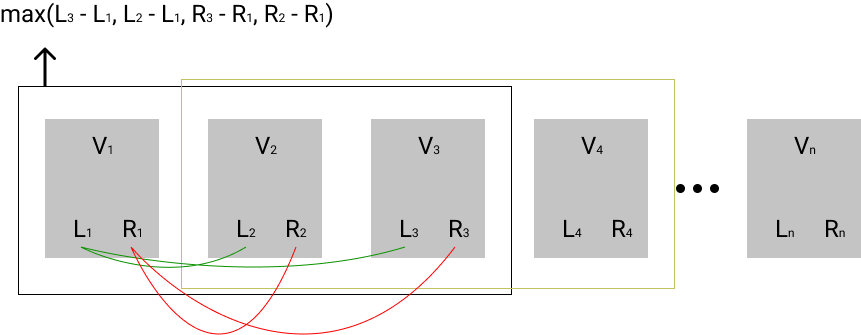

## 6.13 M

Table equivalent to table 1 in the results, describing model score only on the subset of patients who had HbA1c score or disease duration record.

|  | **Image- only HbA1c** | **Patient- only HbA1c** | **Image- only disease duration** | **Patient- only disease duration** |
| --- | --- | --- | --- | --- |
| Mild DR to mtmDR | 0.67 (0.63, 0.71) | 0.69 (0.60, 0.77) | 0.64 (0.59, 0.67) | 0.66 (0.58, 0.73) |
| No DR to mtmDR | 0.65 (0.64, 0.67) | 0.69 (0.66, 0.73) | 0.67 (0.65, 0.68) | 0.71 (0.67, 0.74) |
| mtmDR- to mtmDR+ | 0.74 (0.72, 0.76) | 0.80 (0.76, 0.84) | 0.75 (0.73, 0.77) | 0.81 (0.78, 0.84) |
| Any DR Progression | 0.69 (0.68, 0.71) | 0.75 (0.71, 0.78) | 0.70 (0.69, 0.72) | 0.75 (0.72, 0.78) |

## 6.14 N

Testing the model on patients with different ages.

| **Patient age** | **By image** | **By patient** |
| --- | --- | --- |
| **20-29** | 0.86 (0.79, 0.91) | 0.92 (0.74, 0.98) |
| **30-39** | 0.81 (0.76, 0.86) | 0.87 (0.77, 0.94) |
| **40-49** | 0.73 (0.70, 0.77) | 0.79 (0.71, 0.85) |
| **50-59** | 0.76 (0.74, 0.78) | 0.82 (0.77, 0.86) |
| **60-69** | 0.69 (0.66, 0.72) | 0.75 (0.68, 0.80) |

## 6.15 O

Testing the model on images taken using different camera types.

| **Device** | **Image Number** | **AUC** |
| --- | --- | --- |
| **Canon CR2** | 3994 | 0.711 (0.686, 0.735) |
| **Centervue DRS** | 2600 | 0.751 (0.712, 0.786) |
| **Topcon NW400** | 8719 | 0.733 (0.712, 0.754) |

| **Device** | **Patient Number** | **AUC** |
| --- | --- | --- |
| **device** |  |  |
| **Canon CR2** | 652 | 0.765 (0.707, 0.812) |
| **Centervue DRS** | 462 | 0.823 (0.737, 0.883) |
| **Topcon NW400** | 1503 | 0.791 (0.747, 0.829) |

## 6.16 P

Testing the model on patients with different ethnicities

| **Ethnicity** | **Image Number** | **AUC** |
| --- | --- | --- |
| **African Descent** | 1315 | 0.723 (0.664, 0.777) |
| **Asian** | 912 | 0.728 (0.664, 0.782) |
| **Caucasian** | 1181 | 0.716 (0.649, 0.773) |
| **Indian subcontinent origin** | 458 | 0.749 (0.673, 0.809) |
| **Latin American** | 9484 | 0.733 (0.715, 0.750) |
| **ethnicity not specified** | 1408 | 0.774 (0.719, 0.820) |

| **Ethnicity** | **Patient Number** | **AUC** |
| --- | --- | --- |
| **African Descent** | 237 | 0.782 (0.640, 0.873) |
| **Asian** | 155 | 0.782 (0.644, 0.892) |
| **Caucasian** | 205 | 0.796 (0.622, 0.895) |
| **Indian subcontinent origin** | 75 | 0.786 (0.584, 0.895) |
| **Latin American** | 1656 | 0.791 (0.753, 0.826) |
| **ethnicity not specified** | 246 | 0.820 (0.702, 0.900) |
